# Reported Cases of Multisystem Inflammatory Syndrome in Children (MIS-C) Aged 12–20 Years in the United States Who Received COVID-19 Vaccine, December 2020 through August 2021

**DOI:** 10.1101/2022.01.03.22268681

**Authors:** Anna R. Yousaf, Margaret M. Cortese, Allan W. Taylor, Karen R. Broder, Matthew E. Oster, Joshua M. Wong, Alice Y. Guh, David W. McCormick, Satoshi Kamidani, Elizabeth P. Schlaudecker, Kathryn Edwards, C. Buddy Creech, Mary A. Staat, Ermias D. Belay, Paige Marquez, John R. Su, Mark B. Salzman, Deborah Thompson, Angela P. Campbell, the MIS-C Investigation Authorship Group

## Abstract

**Background:** Multisystem inflammatory syndrome in children (MIS-C) is a hyperinflammatory condition associated with antecedent SARS-CoV-2 infection. In the United States, reporting of MIS-C after vaccination is required under COVID-19 vaccine emergency use authorizations. This case series describes persons aged 12–20 years with MIS-C following COVID-19 vaccination reported to passive surveillance systems or through clinician outreach to CDC.

**Methods:** We investigated potential cases of MIS-C after COVID-19 vaccination reported to CDC’s health department-based national MIS-C surveillance, the Vaccine Adverse Event Reporting System (VAERS, co-administered by CDC and the U.S. FDA), and CDC’s Clinical Immunization Safety Assessment Project (CISA) from December 14, 2020, to August 31, 2021. We describe cases meeting the CDC MIS-C case definition. Any positive SARS-CoV-2 serology test satisfied the case criteria although anti-nucleocapsid antibody indicates SARS-CoV-2 infection, while anti-spike protein antibody indicates either infection or COVID-19 vaccination.

**Findings:** We identified 21 persons with MIS-C after COVID-19 vaccination. Of these 21 persons, median age was 16 years (range, 12–20 years); 13 (62%) were male. All were hospitalized; 12 (57%) had intensive care unit admission, and all were discharged home. Fifteen (71%) of the 21 had laboratory evidence of past or recent SARS-CoV-2 infection, and six (29%) did not. Through August 2021, 21,335,331 persons aged 12–20 years had received ≥1 dose of COVID-19 vaccine, making the overall reporting rate for MIS-C following vaccination 1·0 case per million persons receiving ≥1 vaccine dose in this age group. The reporting rate for those without evidence of SARS-CoV-2 infection was 0·3 cases per million vaccinated persons.

**Interpretation:** In our case series, we describe a small number of persons with MIS-C who had received ≥1 COVID-19 vaccine dose before illness onset. Continued reporting of potential cases and surveillance for MIS-C illnesses after COVID-19 vaccination is warranted.

**Funding:** This work was supported by the Centers for Disease Control and Prevention Clinical Immunization Safety Assessment (CISA] Project contracts 200-2012-50430-0005 to Vanderbilt University Medical Center and 200-2012-53661 to Cincinnati Children’s Hospital Medical Center.

**Research in context panel:** *Evidence before this study:* Multisystem inflammatory syndrome in children (MIS-C), also known as paediatric inflammatory multisystem syndrome temporally associated with SARS-CoV-2 (PIMS-TS), is an uncommon, but serious, complication described after SARS-CoV-2 infection that is characterized by a generalized hyperinflammatory response. A review of the literature using PubMed identified reports of six persons aged 12–20 years who developed MIS-C following COVID-19 vaccination. Search terms used to identify these reports were: “multisystem inflammatory syndrome in children”, “MIS-C”, “MISC”, “multisystem inflammatory syndrome in adults”, “MIS-A”, “MISA”, “paediatric inflammatory multisystem syndrome”, and “PIMS-TS” each with any COVID-19 vaccine type. There were no exclusion criteria (i.e., all ages and languages).

*Added value of this study:* We conducted integrated surveillance for MIS-C after COVID-19 vaccination using two passive surveillance systems, CDC’s MIS-C national surveillance and the Vaccine Adverse Event Reporting System (VAERS), and clinician or health department outreach to CDC, including through Clinical Immunization Safety Assessment (CISA) Project consultations. We investigated reports of potential MIS-C occurring from December 14, 2020, to August 31, 2021, in persons aged 12–20 years any time after receipt of COVID-19 vaccine to identify those that met the CDC MIS-C case definition. Any positive serology test was accepted as meeting the CDC MIS-C case definition, although anti-nucleocapsid antibody is indicative of SARS-CoV-2 infection, while anti-spike protein antibody may be induced either by SARS-CoV-2 infection or by COVID-19 vaccination. We investigated 47 reports and identified 21 persons with MIS-C after receipt of COVID-19 vaccine. Of the 21 persons with MIS-C, median age was 16 years (range 12–20 years), and 13 (62%) were male. Fifteen (71%) had laboratory evidence of past or recent SARS-CoV-2 infection (positive SARS-CoV-2 nucleic acid amplification test [NAAT], viral antigen, or serology test before or during MIS-C illness evaluation), and 5 (33%) of those 15 had illness onset after their second vaccine dose. Six (29%) of 21 persons had no laboratory evidence of past or recent SARS-CoV-2 infection, and five of those six (83%) had onset of MIS-C after the second vaccine dose.

*Implications of all the available evidence:* During the first nine months of the COVID-19 vaccination program in the United States, >21 million persons aged 12 to 20 years received ≥1 dose of COVID-19 vaccine as of August 31, 2021. This case series describes MIS-C in 21 persons following vaccine receipt during this time period; the majority of persons reported also had evidence of SARS-CoV-2 infection. The surveillance has limitations, but our findings suggest that MIS-C as identified in this report following COVID-19 vaccination is rare. In evaluating persons with a clinical presentation consistent with MIS-C after COVID-19 vaccination it is important to consider alternative diagnoses, and anti-nucleocapsid antibody testing may be helpful. Continued surveillance for MIS-C illness after COVID-19 vaccination is warranted, especially as pediatric COVID-19 vaccination expands. Providers are encouraged to report potential MIS-C cases after COVID-19 vaccination to VAERS.

## Introduction

Multisystem inflammatory syndrome in children (MIS-C), also known as paediatric inflammatory multisystem syndrome temporally associated with SARS-CoV-2 (PIMS-TS), is a rare but serious complication of SARS-CoV-2 infection in children and adolescents that generally occurs 2–6 weeks after SARS-CoV-2 infection.^1, 2^ MIS-C, first recognized in April, 2020, is characterized by fever, systemic inflammation, shock, and multisystem organ involvement.^1-6^ From May 14, 2020, through November 30, 2021, 5,973 cases were reported to the CDC’s MIS-C national surveillance system.^7^ The pathogenesis of MIS-C is hypothesized to involve a dysregulated immune response to SARS-CoV-2 infection; host genetics may alter susceptibility to developing MIS-C.^8-11^

CDC and the Food and Drug Administration (FDA) included MIS-C on a list of adverse events of special interest (AESI) for COVID-19 vaccine safety monitoring after emergency use authorization of COVID-19 vaccines in the United States, because of its known association with SARS-CoV-2 infection.^12, 13^ All COVID-19 vaccines currently authorized for use in the United States require reporting of this condition after COVID-19 vaccination.^14^ International vaccine and pharmacovigilance experts have also supported the need for close monitoring of MIS-C after COVID-19 vaccination.^15, 16^

Conducting surveillance for MIS-C after COVID-19 vaccination is challenging because MIS-C in general is a difficult diagnosis to make as it has no specific biomarker and may resemble other disease processes, including acute COVID-19 infection, Kawasaki disease, and toxic shock syndrome.^1-5^ Also, with wide circulation of SARS-CoV-2 occurring concurrently with administration of millions of COVID-19 vaccine doses, some cases of MIS-C caused by SARS-CoV-2 infections acquired before vaccine protection are expected to occur by chance alone and will appear to be temporally associated with vaccine; after a full vaccine series, some MIS-C cases from SARS-CoV-2 infection may occur as protection against infection is <100%.

Recognizing these challenges, we conducted integrated surveillance for MIS-C after COVID-19 vaccination using two passive surveillance systems: CDC’s MIS-C national surveillance system and the Vaccine Adverse Event Reporting System (VAERS), and clinician or health department outreach to CDC, including through Clinical Immunization Safety Assessment (CISA) Project consultations.^7, 17-19^

We investigated reports of potential MIS-C in persons aged 12–20 years who had previously received COVID-19 vaccine during the initial months of the U.S. COVID-19 vaccination program, a period when there was widespread SARS-CoV-2 circulation. We aimed to describe the demographic and clinical features of MIS-C after COVID-19 vaccination, including information on past SARS-CoV-2 infection and COVID-19 vaccination.

## Methods

We identified potential MIS-C cases occurring any time after COVID-19 vaccination in persons aged 12-20 years at time of MIS-C illness onset through CDC’s MIS-C national surveillance system, VAERS, and clinician or health department outreach to CDC and the CISA Project.^7, 17-19^ We investigated reports to determine if the illnesses met the CDC MIS-C case definition. The CDC MIS-C case definition requires fever, hospitalization with an illness with multisystem organ involvement, laboratory evidence of inflammation, and a positive SARS-CoV-2 reverse transcription-polymerase chain reaction (RT-PCR), viral antigen, or serology test or recent exposure to a confirmed COVID-19 case (Figure 1).^6^ This definition was published before COVID-19 vaccination authorization. While anti-nucleocapsid antibody is indicative of prior or current SARS-CoV-2 infection, anti-spike protein antibody may be induced either by SARS-CoV-2 infection or by COVID-19 vaccination (including the three vaccines authorized in the United States).^14^ A positive anti-spike or anti-nucleocapsid antibody can be used to satisfy the SARS-CoV-2 test criterion of the CDC case definition.

**Figure 1.**
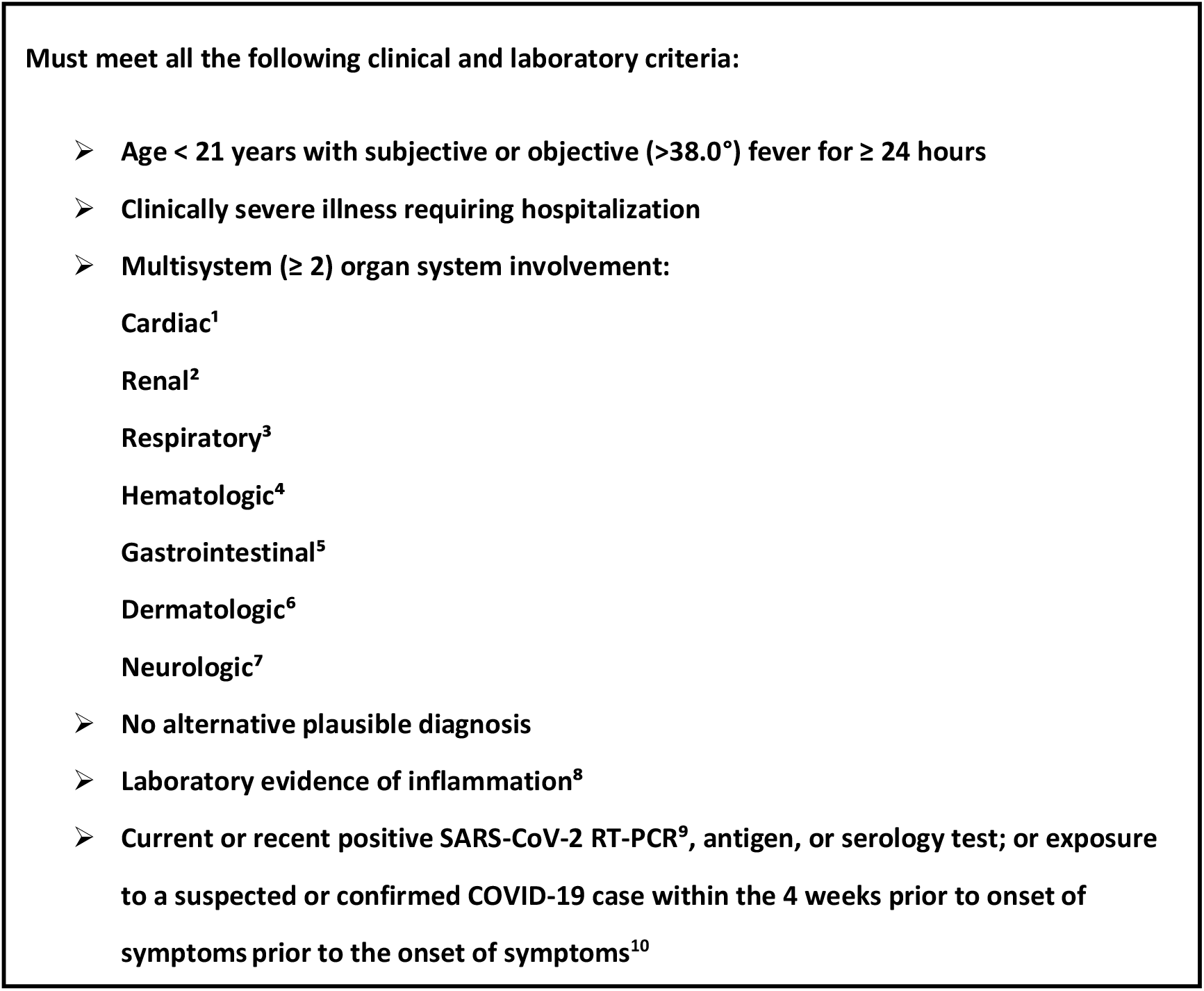
Centers for Disease Control and Prevention multisystem inflammatory syndrome in children (MIS-C) case definition^6^ ^1^Cardiac involvement includes elevated troponin, elevated B-type natriuretic peptide (BNP)/ N-terminal pro hormone BNP (NT-proBNP), arrythmia, coronary artery aneurysm, cardiac dysfunction, or shock; ^2^Renal involvement includes acute kidney injury or renal failure; ^3^Respiratory involvement includes pneumonia, acute respiratory distress syndrome (ARDS), pleural effusion; ^4^Hematologic involvement includes elevated D-dimer, thrombophilia, thrombocytopenia; ^5^Gastrointestinal involvement includes elevated bilirubin, elevated liver enzymes, diarrhea; ^6^Dermatologic involvement includes rash or mucocutaneous lesions; ^7^Neurologic involvement includes cerebrovascular accident, aseptic meningitis encephalopathy, or headache; ^8^Laboratory evidence of inflammation includes elevated C-reactive protein (CRP), erythrocyte sedimentation rate (ESR), fibrinogen, procalcitonin, d-dimer, ferritin, lactic acid dehydrogenase (LDH), interleukin 6 (IL-6), or neutrophils, or reduced lymphocytes or albumin; ^9^For this investigation, this criterion could be satisfied by any type of nucleic acid amplification test (NAAT); ^10^The exposure criterion was not used in this investigation

CDC’s national MIS-C surveillance is a passive reporting system in which health departments voluntarily report cases of MIS-C; collection of COVID-19 vaccination status began May 21, 2021.^7^ We queried the national MIS-C surveillance system twice weekly for persons with MIS-C illness onset date after vaccination date. We reviewed reports weekly of persons with MIS-C onset after COVID-19 vaccination made to VAERS, a passive national surveillance system for vaccine adverse events jointly managed by CDC and FDA which receives spontaneous reports from healthcare providers, health departments, vaccine manufacturers, and the public.^18, 19^ We searched VAERS weekly for reports with coding or free text mention of possible multisystem inflammation or MIS-C (Supplemental Table 1). Additionally, clinicians at CDC and FDA reviewing selected VAERS reports as part of COVID-19 surveillance vaccine safety surveillance referred reports of potential MIS-C to our team for further review. We also received notification of potential MIS-C cases if a provider contacted the MIS-C national surveillance team or requested a CISA consultation.^17^ We encouraged reporting to VAERS for cases first detected by MIS-C national surveillance or through outreach to CDC. We did not specify a maximum interval from COVID-19 vaccination to illness onset in any system searches.

A multidisciplinary team consisting of clinical and surveillance staff from CDC, FDA, and investigators from CDC’s CISA Project adjudicated cases together at least twice monthly. CDC physicians reviewed medical records and presented case summaries to the team. Some potential cases were also discussed with the treating clinicians and health department officials when additional information was needed for adjudication. The investigation team sometimes provided suggestions for laboratory testing; however, clinical evaluation of alternative diagnoses was at the discretion of the treating clinicians. Persons in whom the study team considered myocarditis as a plausible alternative diagnosis based on clinical judgment received additional review with CISA cardiologists to differentiate between myocarditis and cardiac manifestations of MIS-C.^20^

Persons with potential MIS-C after vaccination were classified into two groups: 1) meeting the CDC MIS-C case definition, or 2) not meeting for the CDC MIS-C case definition. Persons with MIS-C were further stratified by laboratory evidence of past or recent SARS-CoV-2 infection. Laboratory evidence of infection was defined as any positive SARS-CoV-2 nucleic acid amplification test (NAAT), including RT-PCR, or viral antigen test before or during MIS-C illness evaluation, or a positive anti-nucleocapsid antibody test during MIS-C illness evaluation. Persons were classified as having no evidence of SARS-CoV-2 infection if they satisfied all of the following: 1) no known history of a positive SARS-CoV-2 test before MIS-C illness onset, 2) a negative SARS-CoV-2 NAAT and/or viral antigen test during MIS-C illness evaluation, and 3) a negative anti-nucleocapsid antibody test during MIS-C illness evaluation. Persons who met these criteria and tested positive for anti-spike antibody were also considered to have no laboratory evidence of SARS-CoV-2 infection as anti-spike antibody was presumed to be vaccine-derived. All potential cases were also assessed using the case definition of the Brighton Collaboration for MIS-C cases; this definition differs from the CDC MIS-C case definition in that it uses a tiered approach to diagnostic certainty and uniquely includes COVID-19 vaccination status as a criterion.^15^

We collected demographic and clinical features of persons with MIS-C from records provided by reporting providers. We defined clinical phenotypes using CDC MIS-C organ system involvement criteria (Figure 1). We stratified temporal elements such as time from prior infection and time from most recent COVID-19 vaccine dose to illness onset by laboratory evidence of SARS-CoV-2 infection and number of COVID-19 vaccine doses. We described the same characteristics for persons with illness not meeting the case definition because of lack of a positive SARS-CoV-2 test (e.g., NAAT negative, anti-nucleocapsid antibody test negative, and anti-spike antibody test not obtained). These persons would presumably have met the case definition had anti-spike antibody been obtained due to presence of vaccine derived anti-spike antibody. We summarized illness features and possible etiologies for persons not meeting the MIS-C case definition because of the presence of a plausible alternative diagnosis.

We obtained number of vaccine doses administered to persons in the United States aged 12–20 years and number of persons in this age group who had received ≥1 dose through August 31, 2021 from CDC national vaccine surveillance data.^21^

This activity was determined by CDC to meet criteria for public health surveillance as defined in 45 CFR §46.102(I)(2). To protect patient privacy, we present persons by age group, and present some demographic and clinical details in aggregate.

## Results

From December 14, 2020, to August 31, 2021, after deduplication, we identified 62 reports of persons with potential MIS-C who had received a COVID-19 vaccine (Figure 2). Thirteen reports were excluded based on information in the VAERS report alone; records were unavailable to fully investigate two reports (Figure 2), and 47 reports were fully investigated. Of these 47 reports, 21 (45%) described illness meeting the CDC MIS-C case definition, and 26 (55%) did not. All 21 persons with MIS-C had evaluations for alternate etiologies, but no alternative was deemed as plausible as MIS-C. None had a previous history of MIS-C. Of the 26 persons with illness not meeting the MIS-C case definition, three (12%) met MIS-C clinical and inflammatory criteria and did not have an alternative diagnosis but did not meet the case definition because of lack of a positive SARS-CoV-2 test; they were anti-nucleocapsid antibody negative, NAAT negative, and anti-spike antibody was not obtained (Figure 2, Supplemental Tables 2a, 2b, 2c). An additional 18 (69%) did not meet the case definition because they had an alternative more likely diagnosis, and 5 (19%) did not meet other clinical or inflammatory criteria of the case definition (Supplemental Table 3).

**Figure 2.**
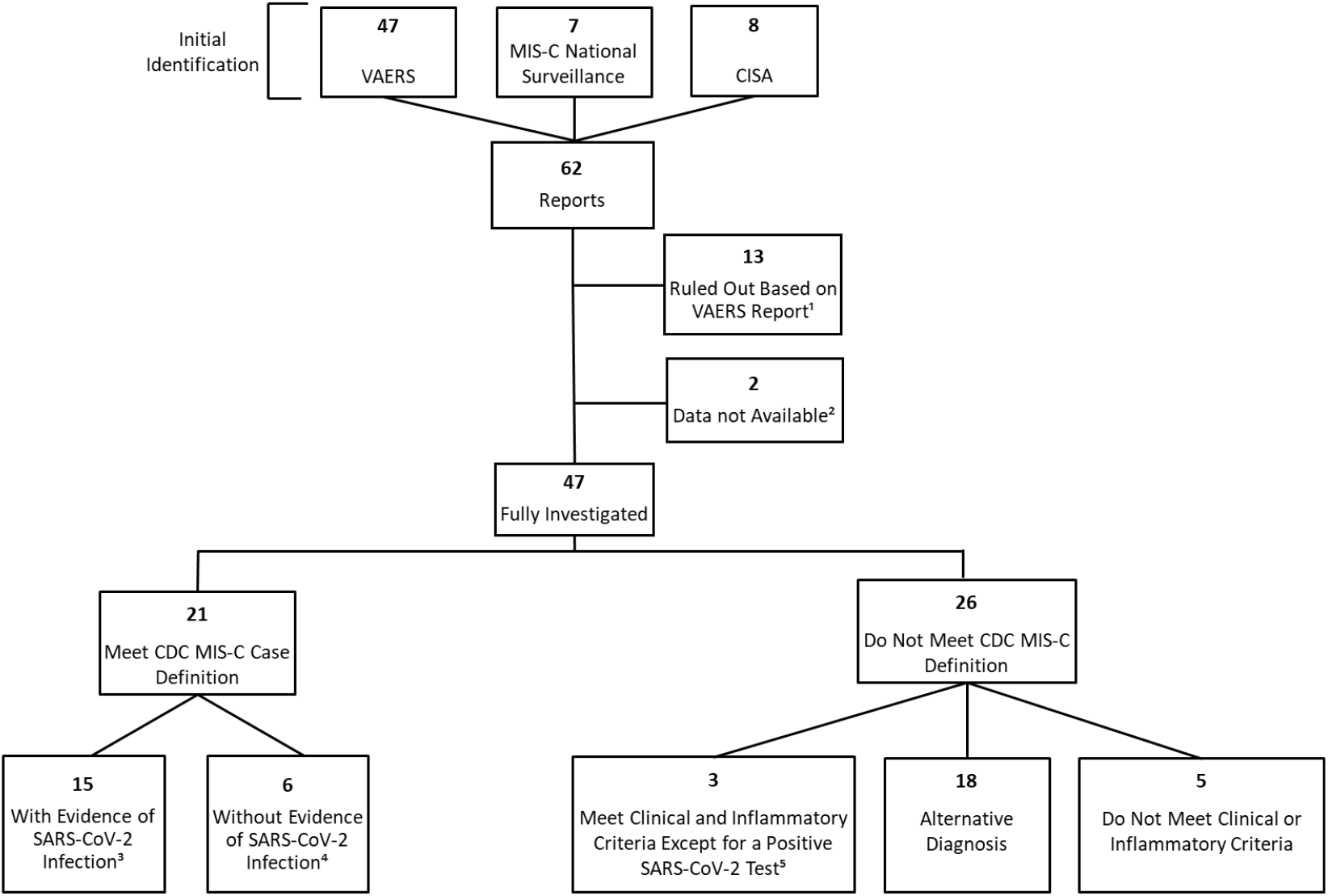
Investigation of potential multisystem inflammatory syndrome in children (MIS-C) in persons who had received COVID-19 vaccination— United States, December 2020 through August 2021 ^1^Reports were ruled out by the VAERS report alone if they were the incorrect age or if MIS-C could be clearly ruled out based on the VAERS report. ^2^Two persons were reported to MIS-C national surveillance but not to VAERS, and medical records have not been obtained. Both had reported MIS-C after 1 dose of COVID-19 vaccine, and both had SARS-CoV-2 nucleic acid amplification test (NAAT) and IgG positive; both clinically improved and were discharged home. ^3^Defined as an illness meeting the CDC MIS-C clinical and inflammatory criteria with a positive NAAT, viral antigen, or anti-nucleocapsid antibody test during or before MIS-C illness evaluation ^4^Defined as an illness meeting the CDC MIS-C clinical and inflammatory criteria with negative NAAT and anti-nucleocapsid antibody tests and a positive anti-spike antibody positive test during MIS-C illness evaluation ^5^Three individuals with an illness after vaccination meeting the CDC MIS-C clinical and inflammatory criteria, a negative anti-nucleocapsid antibody test and negative NAAT test during MIS-C evaluation, and anti-spike antibody test not obtained VAERS: Vaccine Adverse Event Reporting System; CISA: Clinical Immunization Safety Assessment Project; FDA: Food and Drug Administration

As of August 31, 2021, 21,335,331 persons aged 12–20 years had received ≥1 dose of COVID-19 vaccine in the United States: 18,030,614 received Pfizer-BioNTech, 2,603,078 received Moderna, 697,281 received Janssen, and 4,358 did not have manufacturer recorded.^21^

### Persons with MIS-C and laboratory evidence of SARS-CoV-2 infection

Of the 21 persons with MIS-C, 15 (71%) had evidence of SARS-CoV-2 infection (Tables 1, 2). Among these 15 persons with laboratory evidence of infection: ten (67%) were NAAT or viral antigen positive, and five (33%) were anti-nucleocapsid antibody positive and NAAT negative during MIS-C illness with no known positive NAAT or viral antigen test before MIS-C (Table 2). Of those that were NAAT or antigen positive, four had a positive test during MIS-C illness evaluation, one during and also 133 days before MIS-C illness, and five only before MIS-C illness. Of the five persons who were NAAT or viral antigen positive only before MIS-C illness, four were anti-nucleocapsid antibody positive during MIS-C illness, and one was negative (Table 3).

**Table 1.** Demographic characteristics and comorbidities for 21 persons with multisystem inflammatory syndrome in children (MIS-C) following COVID-19 vaccination stratified by laboratory evidence of SARS-CoV-2 infection — United States, December 2020 through August 2021

|  | Total,<br>n=21 (%) | Laboratory Evidence of<br>SARS-CoV-2 Infection,<br>n=15 (%) | No Laboratory Evidence<br>of SARS-CoV-2 Infection,<br>n=6 (%) |
| --- | --- | --- | --- |
| Age at Time of MIS-C Onset (Yrs.) |  |  |  |
| 12–15 | 10 (48) | 7 (47) | 3 (50) |
| 16–17 | 5 (24) | 5 (33) | 0 (0) |
| 18–20 | 6 (29) | 3 (20) | 3 (50) |
| Sex |  |  |  |
| Male | 13 (62) | 10 (67) | 3 (50) |
| Race/Ethnicity |  |  |  |
| Hispanic | 5 (24) | 4 (27) | 1 (17) |
| White, Non-Hispanic | 8 (38) | 4 (27) | 4 (67) |
| Black, Non-Hispanic | 3 (14) | 3 (20) | 0 (0) |
| Asian/Pacific Islander, Non-Hispanic | 3 (14) | 2 (13) | 1 (17) |
| Other race, Non-Hispanic<sup>1</sup> | 2 (10) | 2 (13) | 0 (0) |
| Comorbidities<sup>2</sup> |  |  |  |
| Asthma | 7 (33) | 6 (40) | 1 (17) |
| Obesity | 3 (14) | 2 (13) | 1 (17) |
| Seizure Disorder | 1 (5) | 0 (0) | 1 (17) |

<sup>1</sup>Other race= one with multiple races, one with unknown race
<sup>2</sup>Other conditions listed in medical charts included mental health conditions, gastroesophageal reflux disorder, herniated disc with sciatica.

**Table 2.** SARS-CoV-2 laboratory testing in 21 persons with multisystem inflammatory syndrome in children (MIS-C) following COVID-19 vaccination stratified by number of COVID-19 vaccine doses received before MIS-C illness onset— United States, December 2020 through August 2021

|  |  |  | Total (n) | Onset After COVID-19 Vaccine Dose 1, n (%) | Onset After COVID-19 Vaccine Dose 2, n (%) |
| --- | --- | --- | --- | --- | --- |
| **MIS-C** |  |  | **21** | **10 (42)** | **14 (58)** |
|  | With Evidence of SARS-CoV-2 infection |  | 15 | 10 (67) | 5 (33) |
|  | NAAT positive (past or recent)<sup>1</sup> |  | 10 | 5 (50) | 5 (50) |
|  | NAAT negative and positive anti-nucleocapsid antibody |  | 5 | 4 (80) | 1 (20) |
|  | Without evidence of SARS-CoV-2 infection |  | 6 | 1 (17) | 5 (83) |
|  | NAAT negative, anti- nucleocapsid antibody negative, anti-spike antibody positive<sup>2</sup> |  | 6 | 1 (17) | 5 (83) |

<sup>1</sup>Includes five persons with positive SARS-CoV-2 NAAT tests before MIS-C illness, four persons with positive SARS-CoV-2 NAAT tests during MIS-C illness, and one person with a positive SARS-CoV-2 NAAT test before and during MIS-C illness
<sup>2</sup>Includes NAAT results from during MIS-C illness evaluation; these individuals did not have any reported positive NAAT results before MIS-C illness

**Table 3.**
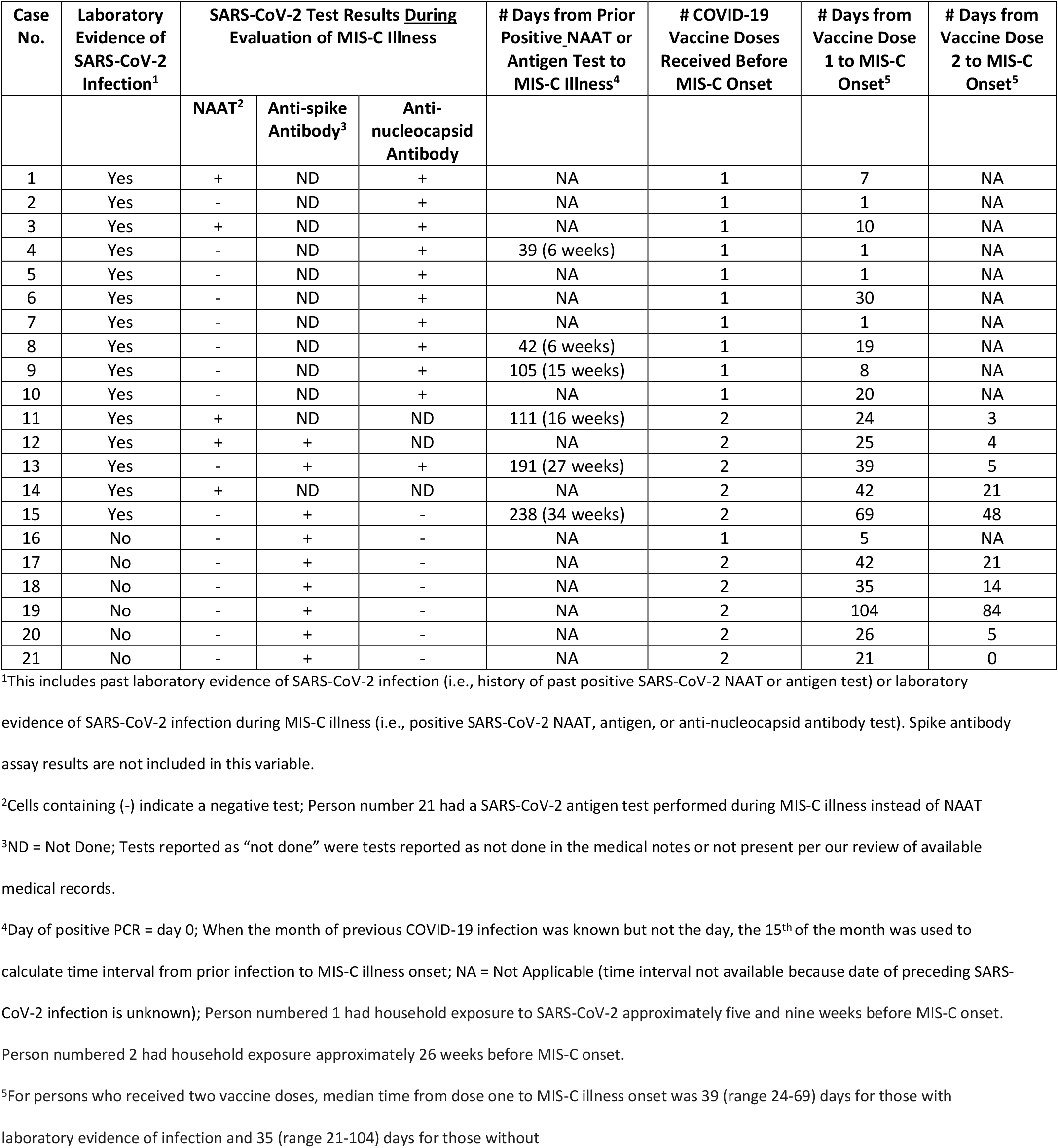
SARS-CoV-2 testing and temporal features of 21 persons with multisystem inflammatory syndrome in children (MIS-C) following COVID-19 vaccination— United States, December 2020 through August 2021

| Case No. | Laboratory Evidence of SARS-CoV-2 Infection<sup>1</sup> | SARS-CoV-2 Test Results During Evaluation of MIS-C Illness |  |  | # Days from Prior Positive NAAT or Antigen Test to MIS-C Illness<sup>4</sup> | # COVID-19 Vaccine Doses Received Before MIS-C Onset | # Days from Vaccine Dose 1 to MIS-C Onset<sup>5</sup> | # Days from Vaccine Dose 2 to MIS-C Onset<sup>5</sup> |
| --- | --- | --- | --- | --- | --- | --- | --- | --- |
|  |  | NAAT<sup>2</sup> | Anti-spike Antibody<sup>3</sup> | Anti-nucleocapsid Antibody |  |  |  |  |
| 1 | Yes | + | ND | + | NA | 1 | 7 | NA |
| 2 | Yes | - | ND | + | NA | 1 | 1 | NA |
| 3 | Yes | + | ND | + | NA | 1 | 10 | NA |
| 4 | Yes | - | ND | + | 39 (6 weeks) | 1 | 1 | NA |
| 5 | Yes | - | ND | + | NA | 1 | 1 | NA |
| 6 | Yes | - | ND | + | NA | 1 | 30 | NA |
| 7 | Yes | - | ND | + | NA | 1 | 1 | NA |
| 8 | Yes | - | ND | + | 42 (6 weeks) | 1 | 19 | NA |
| 9 | Yes | - | ND | + | 105 (15 weeks) | 1 | 8 | NA |
| 10 | Yes | - | ND | + | NA | 1 | 20 | NA |
| 11 | Yes | + | ND | ND | 111 (16 weeks) | 2 | 24 | 3 |
| 12 | Yes | + | + | ND | NA | 2 | 25 | 4 |
| 13 | Yes | - | + | + | 191 (27 weeks) | 2 | 39 | 5 |
| 14 | Yes | + | ND | ND | NA | 2 | 42 | 21 |
| 15 | Yes | - | + | - | 238 (34 weeks) | 2 | 69 | 48 |
| 16 | No | - | + | - | NA | 1 | 5 | NA |
| 17 | No | - | + | - | NA | 2 | 42 | 21 |
| 18 | No | - | + | - | NA | 2 | 35 | 14 |
| 19 | No | - | + | - | NA | 2 | 104 | 84 |
| 20 | No | - | + | - | NA | 2 | 26 | 5 |
| 21 | No | - | + | - | NA | 2 | 21 | 0 |

<sup>1</sup>This includes past laboratory evidence of SARS-CoV-2 infection (i.e., history of past positive SARS-CoV-2 NAAT or antigen test) or laboratory evidence of SARS-CoV-2 infection during MIS-C illness (i.e., positive SARS-CoV-2 NAAT, antigen, or anti-nucleocapsid antibody test). Spike antibody assay results are not included in this variable.
<sup>2</sup>Cells containing (-) indicate a negative test; Person number 21 had a SARS-CoV-2 antigen test performed during MIS-C illness instead of NAAT
<sup>3</sup>ND = Not Done; Tests reported as “not done” were tests reported as not done in the medical notes or not present per our review of available medical records.
<sup>4</sup>Day of positive PCR = day 0; When the month of previous COVID-19 infection was known but not the day, the 15<sup>th</sup> of the month was used to calculate time interval from prior infection to MIS-C illness onset; NA = Not Applicable (time interval not available because date of preceding SARS-CoV-2 infection is unknown); Person numbered 1 had household exposure to SARS-CoV-2 approximately five and nine weeks before MIS-C onset. Person numbered 2 had household exposure approximately 26 weeks before MIS-C onset.
<sup>5</sup>For persons who received two vaccine doses, median time from dose one to MIS-C illness onset was 39 (range 24-69) days for those with laboratory evidence of infection and 35 (range 21-104) days for those without

Seven (47%) were aged 12–15 years, five (33%) aged 16–17 years, and three (20%) aged 18–20 years (Table 1). Ten (67%) were male. Four (27%) each were Hispanic and White Non-Hispanic persons and three (20%) were Black Non-Hispanic. The following organ systems were most commonly involved during MIS-C illness: 14 (93%) gastrointestinal, 13 (87%) hematologic, and 13 (87%) cardiac (Supplemental Table 4). Upon assessment using the Brighton case definition, 12 (80%) were definitive or probable cases and three (20%) would not be considered cases (Supplemental Table 4).

All 15 persons with laboratory evidence of SARS-CoV-2 infection had received Pfizer-BioNTech COVID-19 vaccine (the only U.S. COVID-19 vaccine authorized for use in persons aged <18 years during our surveillance), with 10 (66%) receiving one dose and five (33%) receiving two doses before MIS-C illness onset (Table 2). Median time from most recent vaccine dose to MIS-C onset was eight (range 1– 30) days for those who had received only one dose, and five (range 3–48) days for those who had received two doses (Table 3, Supplemental Figure 1).

During their MIS-C hospitalization, 13 (87%) persons were treated with intravenous immunoglobulin (IVIG), 12 (80%) with systemic steroids, and five (33%) with an immune modulator (Table 4). Eight (53%) were admitted to the intensive care unit (ICU). Median length of hospital stay was seven (range 2–20) days. All 15 persons clinically improved and were discharged home.

**Table 4.** Treatment and outcomes for 21 persons with multisystem inflammatory syndrome in children (MIS-C) following COVID-19 vaccination stratified by laboratory evidence of SARS-CoV-2 infection— United States, December 2020 through August 2021

|  | Total,<br>n=21 (%) | Laboratory Evidence of<br>SARS-CoV-2 Infection,<br>n=15 (%) | No Laboratory Evidence<br>of SARS-CoV-2 Infection,<br>n=6 (%) |
| --- | --- | --- | --- |
| Inpatient MIS-C Treatment |  |  |  |
| Intravenous immunoglobulin (IVIG) | 17 (81) | 13 (87) | 4 (67) |
| Systemic steroids | 16 (76) | 12 (80) | 4 (67) |
| Immune modulators | 5 (23) | 5 (33)<sup>1</sup> | 0 (0) |
| Remdesivir | 1 (5) | 1 (7)<sup>2</sup> | 0 (0) |
| None | 3 (14) | 1 (7) | 2 (33) |
| Admitted to intensive care unit | 12 (57) | 8 (53) | 4 (67) |
| Vasopressors | 8 (38) | 6 (40) | 2 (33) |
| Invasive mechanical ventilation | 3 (14) | 2 (13) | 1 (17) |
| Length of hospitalization in days, median (range) | 6 (2–21) | 7 (2–20) | 6 (3–7) |
| Discharged Home | 21 (100) | 15 (100) | 6 (100) |
| Discharge MIS-C Medications |  |  |  |
| Systemic steroids | 15 (71) | 11 (73) | 4 (67) |
| Aspirin | 14 (67) | 10 (67) | 4 (67) |
| Enoxaparin | 2 (10) | 2 (13) | 0 (0) |
| Angiotensin-converting enzyme inhibitors | 2 (10) | 2 (13) | 0 (0) |
| Other<sup>3</sup> | 2 (10) | 2 (13) | 0 (0) |
| None | 4 (19) | 2 (13) | 2 (33) |

<sup>1</sup>Immune modulators: three persons treated with anakinra; two persons treated with infliximab
<sup>2</sup>In addition, one person (not counted here) with a positive SARS-CoV-2 NAAT during MIS-C illness received an unspecified anti-viral medication
<sup>3</sup>Other medication: anakinra; furosemide, metoprolol

### Persons with MIS-C without laboratory evidence of SARS-CoV-2 infection

Of the 21 persons with MIS-C, six (29%) had positive anti-spike antibody testing only (Tables 1, 2) and were classified as not having laboratory evidence of SARS-CoV-2 infection (Figure 2). None of these persons had a history of a positive SARS-CoV-2 test before MIS-C illness, and all had negative SARS-CoV-2 NAAT and anti-nucleocapsid antibody tests during MIS-C illness (Table 3).

Of the six, three (50%) were aged 12–15 years and three (50%) aged 18–20 years (Table 1). Three (50%) were male; four (67%) were White Non-Hispanic (Table 1). These persons presented with varied organ system involvement: six (100%) cardiac (including two with shock), and five (83%) hematologic (Supplemental Table 5). Applying the Brighton case definition, all were definitive or probable cases (Supplemental Table 5).

All had received Pfizer-BioNTech vaccine; one person (11%) received only one dose five days before MIS-C onset, and five (83%) received two doses before MIS-C illness onset (Table 2). Median time from vaccination with dose 2 to MIS-C onset was 14 (range 0–84) days for those who received two doses (Table 3, Supplemental Figure 2).

Four (67%) persons were treated for MIS-C with IVIG, four (67%) with systemic steroids (Table 4). Median length of stay was six days (range 3–7) days. Four (67%) were admitted to the ICU; all six clinically improved and were discharged home.

## Discussion

As part of the United States’ comprehensive efforts to monitor COVID-19 vaccine safety following authorization, we investigated reported potential U.S. MIS-C cases during a period of widespread SARS-CoV-2 circulation. In this case series, we describe 21 persons with illness meeting the CDC MIS-C case definition. All received the Pfizer-BioNTech vaccine, consistent with the age eligibility of COVID-19 vaccines during the investigation period. Most cases had laboratory evidence of SARS-CoV-2 infection, although prior positive NAAT in three persons occurred outside the typical time frame for MIS-C (105, 191, and 238 days before illness onset), and one other had household exposure outside the typical period. In four others, there was no known exposure to inform the timing of infection that resulted in their anti-nucleocapsid antibody positivity. Overall, the reporting rate for MIS-C through August 2021, was 21/21,335,331, or 1·0 cases per million persons aged 12–20 years who had received ≥1 dose of any COVID-19 vaccine. Although there is not a direct comparator background rate, the reporting rate of illness meeting the MIS-C definition in persons who had received COVID-19 vaccine is substantially lower than the previously published incidence of MIS-C among unvaccinated persons who had SARS-CoV-2 infection. Using a denominator of SARS-CoV-2 infections among unvaccinated persons, a previous study estimated an adjusted incidence of MIS-C from April to June 2020 of 224 (95% CI, 160– 312) in children aged 11–15 years and 164 (95% CI 110–243) in those aged 16–20 years per million SARS-CoV-2 infections.^22^

We identified six cases of MIS-C that occurred following COVID-19 vaccination without evidence of SARS-CoV-2 infection, a reporting rate of 6/21,335,331, or 0.3 cases per million persons aged 12–20 years who had received ≥1 dose of COVID-19 vaccine. It has been hypothesized that a dysregulated immune response associated with SARS-CoV-2 infection may potentially also be associated with exposure to COVID-19 vaccine.^10, 15, 23^As with our cases who had evidence of prior infection outside of the usual period for development of MIS-C, the contribution of vaccination, if any, to the illnesses in cases without evidence of infection is unknown and cannot be determined with our surveillance data. It is possible some of these six cases had other unrecognized inflammatory conditions. Because the pre-pandemic background incidence of illnesses with unidentified etiology that would meet the clinical criteria of the MIS-C case definition is unknown, we cannot estimate how often such illnesses would be expected to occur temporally associated with vaccine by chance alone. In addition, given the limitations of laboratory assays and detection sensitivities of each test, some of the six cases may have been infected with SARS-CoV-2 in the recent past, and vaccination may be coincidental to the subsequent MIS-C illness. Children often have unrecognized SARS-CoV-2 infection associated with mild or absent symptoms.^3, 5^ Persons with mild or asymptomatic illness may be less likely to generate anti-nucleocapsid antibody, and anti-nucleocapsid antibody from prior infection wanes over time, particularly in those with mild infection.^8, 24, 25^ These limitations could lead to misclassification of SARS-CoV-2 infection status. In addition to the six persons without evidence of SARS-CoV-2 infection, we identified three others who met clinical and inflammatory criteria and did not have evidence of SARS-CoV-2 infection but did not meet MIS-C case definition because anti-spike antibody was not obtained.

Neither MIS-C nor multisystem inflammatory syndrome in adults (MIS-A) was reported in the clinical trials of COVID-19 vaccines used in the United States.^14, 26^ Globally, MIS-C following COVID-19 vaccination has been reported in the literature for six persons <21 years of age.^23, 27-30^ Two of these persons are from the United States and are included in our case series; both had evidence of SARS-CoV-2 infection.^23^ A third reported person from the United States had received dose two of the Pfizer-BioNTech vaccine two months prior to presentation and had evidence of infection with a positive anti-nucleocapsid antibody test during MIS-C illness evaluation (NAAT was negative).^30^ Two other reported cases were European adolescents; one without evidence of SARS-CoV-2 infection (onset five days after Pfizer-BioNTech dose 2), the other SARS-CoV-2 NAAT negative and anti-nucleocapsid antibody testing not described (onset 10 weeks after Pfizer-BioNTech dose 2).^27, 28^ Lastly, a sixth report in the literature was of a 12-year-old child from Saudi Arabia who developed MIS-C five weeks after a dose of Moderna COVID-19 vaccine (he received a dose of Pfizer-BioNTech vaccine three weeks before the Moderna dose); this child had negative SARS-CoV-2 NAAT testing, anti-spike antibody testing was positive, and anti-nucleocapsid antibody testing was not performed.^29^ Several adults have also been reported with MIS-A following COVID-19 vaccination, most of whom had evidence of previous SARS-CoV-2 infection.^23,26^

This investigation highlights the challenges of diagnosing MIS-C and importance of a thorough clinical evaluation. While the CDC MIS-C case definition can be met with any type of SARS-CoV-2 antibody test (anti-spike, anti-nucleocapsid, or undifferentiated), testing for anti-nucleocapsid antibody in persons with suspected MIS-C after COVID-19 vaccination may differentiate between vaccine-versus infection-derived antibody. As noted above, however, antibody titers can wane over time. Conversely, as cumulatively higher rates of SARS-CoV-2 infection result in higher anti-nucleocapsid seroprevalence among children, without a test available to indicate how recently SARS-CoV-2 infection occurred, detection of anti-nucleocapsid antibody may be coincidental and may not distinguish MIS-C from other clinically similar syndromes (e.g., toxic shock syndrome). Therefore, a thorough clinical evaluation to elucidate alternative etiologies is important, as many conditions can mimic MIS-C (Supplemental Table 3).

These data are subject to additional limitations. The national MIS-C and VAERS surveillance platforms are both passive reporting systems, and it is likely there is under-reporting, particularly since receipt of vaccine is not part of the MIS-C case definition. Vaccinated persons being evaluated who have negative SARS-CoV-2 NAAT and anti-nucleocapsid serology tests (and do not have anti-spike antibody testing performed) might not be reported since they do not satisfy the CDC case definition and might not be suspected of having MIS-C. We identified three such patients with our surveillance procedures and acknowledge such patients would be incompletely captured. For practicality, we used a limited number of specific terms for our search strategy in the VAERS component of our surveillance. However, our VAERS text search strategy does not depend on reports listing a final diagnosis of MIS-C. Time from vaccine receipt to MIS-C onset may be underestimated for the subset that had shorter intervals because fever and headache are both common short-term vaccine reactogenicity events after COVID-19 vaccination and also symptoms of MIS-C. Clinical and laboratory workup was not standardized across cases, and although cases were adjudicated by an interdisciplinary team, there is no definitive diagnostic test to confirm MIS-C.

In conclusion, during the first nine months of the COVID-19 vaccination program in the United States, a period when SARS-CoV-2 was widely circulating, we identified a small number of persons aged 12-20 years with MIS-C after COVID-19 vaccination; most had laboratory evidence of past or recent SARS-CoV-2 infection. The surveillance has limitations, but our findings suggest that MIS-C without evidence of SARS-CoV-2 infection is rare following COVID-19 vaccination (reporting rate <1 case per million vaccinated persons aged 12–20 years). In evaluating persons with an MIS-C clinical presentation after COVID-19 vaccination, it is important to consider alternative diagnoses, and anti-nucleocapsid antibody testing may be helpful. Continued surveillance for MIS-C illness after COVID-19 vaccination is warranted, especially as pediatric COVID-19 vaccination expands, and providers are encouraged to report potential MIS-C cases after COVID-19 vaccination to VAERS at https://vaers.hhs.gov.

## Data Availability

All data produced in the present study are available upon reasonable request to the authors

## Acknowledgements

Michael Melgar, Katherine Lindsay, Allison D. Miller, Michael Wu, Laura D. Zambrano, Theresa Harrington, Narayan Nair, Mike Meltzer, Elaine Miller, Adam Schiller, Brian Kit, Jonathan Soslow, Jeffrey Dendy, The Clinical Immunization Safety Assessment Project, Frank Destefano, Tom Shimabukuro. We would like to thank all local, state, and territorial health departments that contributed MIS-C reports to this investigation. We would like to thank all health care providers who made reports to VAERS and who were involved in the care of the patients described in this investigation.

The findings and conclusions in this report are those of the authors and do not necessarily represent the official position of the Centers for Disease Control and Prevention or the Food and Drug Administration. This work was supported by the Centers for Disease Control and Prevention Clinical Immunization Safety Assessment (CISA] Project contracts 200-2012-50430-0005 to Vanderbilt University Medical Center and 200-2012-53661 to Cincinnati Children’s Hospital Medical Center.

## Contributors

ARY, MMC, AT, KRB, and APC conceived of and designed the study. ARY, AT, and APC wrote the initial draft. ARY, AT, and MMC produced figures and tables. ARY, MMC, AT, JMW, AYG, DWM, SK, PM, OM, MP, JRS, MBS, JO, CL, LEF, MK, KR, KK, BS, and CB helped to procure and clean the data. ARY, AT, MMC, KRB, EDB, APC MEO, EPS, KE, BC, MAS, LMH, DT, SL, and JS adjudicated cases. All authors edited the manuscript, provided feedback on the study, and approved the final manuscript.

## References

1. Feldstein LR, Rose EB, Horwitz SM, et al. Multisystem Inflammatory Syndrome in U.S. Children and Adolescents. N Engl J Med. Jul 23 2020;383(4):334–346. doi:10.1056/NEJMoa2021680

2. Dufort EM, Koumans EH, Chow EJ, et al. Multisystem Inflammatory Syndrome in Children in New York State. N Engl J Med. Jul 23 2020;383(4):347–358. doi:10.1056/NEJMoa2021756

3. Hoste L, Van Paemel R, Haerynck F. Multisystem inflammatory syndrome in children related to COVID-19: a systematic review. Eur J Pediatr. Jul 2021;180(7):2019–2034. doi:10.1007/s00431-021-03993-5

4. Riphagen S, Gomez X, Gonzalez-Martinez C, Wilkinson N, Theocharis P. Hyperinflammatory shock in children during COVID-19 pandemic. Lancet. May 23 2020;395(10237):1607–1608. doi:10.1016/S0140-6736(20)31094-1

5. Feldstein LR, Tenforde MW, Friedman KG, et al. Characteristics and Outcomes of US Children and Adolescents With Multisystem Inflammatory Syndrome in Children (MIS-C) Compared With Severe Acute COVID-19. JAMA. Mar 16 2021;325(11):1074–1087. doi:10.1001/jama.2021.2091

6. Multisystem Inflammatory Syndrome in Children (MIS-C) Associated with Coronavirus Disease 2019 (COVID-19). Health Advisory. CDC Health Alert Network; May 14, 2020. https://emergency.cdc.gov/han/2020/han00432.asp

7. CDC. Health Department-Reported Cases of Multisystem Inflammatory Syndrome in Children (MIS-C) in the United States. Accessed November 8, https://covid.cdc.gov/covid-data-tracker/#mis-national-surveillance

8. Weisberg SP, Connors TJ, Zhu Y, et al. Distinct antibody responses to SARS-CoV-2 in children and adults across the COVID-19 clinical spectrum. Nat Immunol. Jan 2021;22(1):25–31. doi:10.1038/s41590-020-00826-9

9. Rowley AH, Shulman ST, Arditi M. Immune pathogenesis of COVID-19-related multisystem inflammatory syndrome in children. J Clin Invest. Nov 2 2020;130(11):5619–5621. doi:10.1172/JCI143840

10. Chou J, Platt CD, Habiballah S, et al. Mechanisms underlying genetic susceptibility to multisystem inflammatory syndrome in children (MIS-C). J Allergy Clin Immunol. Sep 2021;148(3):732–738 e1. doi:10.1016/j.jaci.2021.06.024

11. Sancho-Shimizu V, Brodin P, Cobat A, et al. SARS-CoV-2-related MIS-C: A key to the viral and genetic causes of Kawasaki disease? J Exp Med. Jun 7 2021;218(6)doi:10.1084/jem.20210446

12. Gubernot D, Jazwa A, Niu M, et al. U.S. Population-Based background incidence rates of medical conditions for use in safety assessment of COVID-19 vaccines. Vaccine. Jun 23 2021;39(28):3666–3677. doi:10.1016/j.vaccine.2021.05.016

13. Nicola Klein JD, Eric, Weintraub ea. Rapid Cycle Analysis (RCA) to monitor the safety of COVID-19 vaccines in near real-time within the Vaccine Safety Datalink. 2021

14. FDA. COVID-19 Vaccines. Accessed November 8. https://www.fda.gov/emergency-preparedness-and-response/coronavirus-disease-2019-covid-19/covid-19-vaccines

15. Vogel TP, Top KA, Karatzios C, et al. Multisystem inflammatory syndrome in children and adults (MIS-C/A): Case definition & guidelines for data collection, analysis, and presentation of immunization safety data. Vaccine. May 21 2021;39(22):3037–3049. doi:10.1016/j.vaccine.2021.01.054

16. European Medicines Agency. Meeting highlights from the Pharmacovigilance Risk Assessment Committee (PRAC) 25-28 October 2021. October 29. Accessed December 21. https://www.ema.europa.eu/en/news/meeting-highlights-pharmacovigilance-risk-assessment-committee-prac-25-28-october-2021

17. CDC. Clinical Immunization Safety Assessment (CISA) Project. Accessed November 8, 2021. https://www.cdc.gov/vaccinesafety/ensuringsafety/monitoring/cisa/index.html

18. Shimabukuro TT, Nguyen M, Martin D, DeStefano F. Safety monitoring in the Vaccine Adverse Event Reporting System (VAERS). Vaccine. Aug 26 2015;33(36):4398–405. doi:10.1016/j.vaccine.2015.07.035

19. Vaccine Adverse Event Reporting System (VAERS) Standard Operating Procedures for COVID-19. Accessed November 8. https://www.cdc.gov/vaccinesafety/pdf/VAERS-v2-SOP.pdf.

20. Patel T, Kelleman M, West Z, et al. Comparison of MIS-C Related Myocarditis, Classic Viral Myocarditis, and COVID-19 Vaccine related Myocarditis in Children. medRxiv. 2021:2021.10.05.21264581. doi:10.1101/2021.10.05.21264581

21. CDC. About COVID-19 Vaccine Delivered and Administration Data. August 30. Accessed November 9. https://www.cdc.gov/coronavirus/2019-ncov/vaccines/distributing/about-vaccine-data.html

22. Payne AB, Gilani Z, Godfred-Cato S, et al. Incidence of Multisystem Inflammatory Syndrome in Children Among US Persons Infected With SARS-CoV-2. JAMA Netw Open. Jun 1 2021;4(6):e2116420. doi:10.1001/jamanetworkopen.2021.16420

23. Salzman MB, Huang CW, O’Brien CM, Castillo RD. Multisystem Inflammatory Syndrome after SARS-CoV-2 Infection and COVID-19 Vaccination. Emerg Infect Dis. May 25 2021;27(7)doi:10.3201/eid2707.210594

24. Ibarrondo FJ, Fulcher JA, Goodman-Meza D, et al. Rapid Decay of Anti-SARS-CoV-2 Antibodies in Persons with Mild Covid-19. N Engl J Med. Sep 10 2020;383(11):1085–1087. doi:10.1056/NEJMc2025179

25. Petersen LR, Sami S, Vuong N, et al. Lack of Antibodies to Severe Acute Respiratory Syndrome Coronavirus 2 (SARS-CoV-2) in a Large Cohort of Previously Infected Persons. Clin Infect Dis. Nov 2 2021;73(9):e3066–e3073. doi:10.1093/cid/ciaa1685

26. Belay ED, Godfred Cato S, Rao AK, et al. Multisystem Inflammatory Syndrome in Adults after SARS-CoV-2 infection and COVID-19 vaccination. Clinical Infectious Diseases. 2021;doi:10.1093/cid/ciab936

27. Chai Q, Nygaard U, Schmidt RC, Zaremba T, Moller AM, Thorvig CM. Multisystem inflammatory syndrome in a male adolescent after his second Pfizer-BioNTech COVID-19 vaccine. Acta Paediatr. Oct 7 2021;doi:10.1111/apa.16141

28. Buchhorn R, Meyer C, Schulze-Forster K, Junker J, Heidecke H. Autoantibody Release in Children after Corona Virus mRNA Vaccination: A Risk Factor of Multisystem Inflammatory Syndrome? Vaccines (Basel). Nov 18 2021;9(11)doi:10.3390/vaccines9111353

29. Abdelgalil AA, Saeedi FA. Multisystem Inflammatory Syndrome in a 12-Year-old Boy After mRNA-SARS-CoV-2 Vaccination. Pediatr Infect Dis J. Dec 21 2021;doi:10.1097/INF.0000000000003442

30. DeJong J, Sainato R, Forouhar M, Robinson D, Kunz A. Multisystem Inflammatory Syndrome in a Previously Vaccinated Adolescent Female With Sickle Cell Disease. Pediatr Infect Dis J. Dec 21 2021;doi:10.1097/INF.0000000000003444

