## Supplemental Materials for "Reported Cases of Multisystem Inflammatory Syndrome in Children (MIS-C) Aged 12–20 Years in the United States Who Received COVID-19 Vaccine, December 2020 through August 2021"

### Supplemental Materials Table of Contents


**Table S1**………………………………………………………………………………………………………………………… 2

**Table S2a, 2b, 2c** ………………………………………………………………………………………………………… 3

**Table S3**………………………………………………………………………………………………………………………… 5

**Table S4**………………………………………………………………………………………………………………………… 7

**Table S5**………………………………………………………………………………………………………………………… 8

**Figure S1**……………………………………………………………………………………………………………………… 9

**Figure S2**……………………………………………………………………………………………………………………… 10

**Supplemental Table 1.** Vaccine Adverse Event System (VAERS) search terms used to identify potential cases of multisystem inflammatory syndrome in children (MIS-C) after receipt of COVID-19 vaccine

| | |
|---|---|
| Medical Dictionary for Regulatory Activities (MedDRA) Preferred Terms | Multisystem Inflammatory Syndrome in Children<br>SIRS (Systemic Inflammatory Response Syndrome) |
| Text String Search Terms | "MIS-C"<br>"MISC"<br>"MIS"<br>"Multisystem Inflammatory Syndrome"<br>"Multisystem inflammatory"<br>"Multisystem inflammation"<br>"Multisystem" + "inflammation"<br>"Multisystem" + "inflammatory" |

**Supplemental Table 2a.** Demographic and clinical features of three persons with illness meeting MIS-C clinical and inflammatory criteria except for a positive SARS-CoV-2 test— United States, December 2020 through August 2021

| | | Total, n=3 (%) |
|---|---|---|
| Age at Time of Illness Onset (Yrs.) | | |
| | 12–15 | 3 (100) |
| Sex | | |
| | Male | 3 (100) |
| Race/Ethnicity | | |
| | Hispanic | 2 (67) |
| | White, Non-Hispanic | 1 (33) |
| Inpatient MIS-C Treatment | | |
| | Intravenous immunoglobulin (IVIG) | 3 (100) |
| | Systemic steroids | 3 (100) |
| | Immune modulators | 0 (0) |
| Admitted to intensive care unit | | 0 (0) |
| Vasopressors | | 0 (0) |
| Invasive mechanical ventilation | | 0 (0) |
| Length of hospitalization in days, median (range) | | 5 (3–9) |
| Discharged Home | | 3 (100) |

**Supplemental Table 2b.** SARS-CoV-2 testing and temporal features of three persons with illness meeting MIS-C clinical and inflammatory criteria except for a positive SARS-CoV-2 test— United States, December 2020 through August 2021

| Case No. | Laboratory Evidence of SARS-CoV-2 Infection[1] | SARS-CoV-2 Test Results During Evaluation of MIS-C Illness[2] | | | # Days from Prior Positive NAAT or Antigen Test to MIS-C Illness | # COVID-19 Vaccine Doses Received Before MIS-C Onset | # Days from Vaccine Dose 1 to MIS-C Onset | # Days from Vaccine Dose 2 to MIS-C Onset |
|---|---|---|---|---|---|---|---|---|
| | | **NAAT** | **Anti-spike Antibody** | **Anti-nucleocapsid Antibody** | | | | |
| 1 | No | - | ND | - | NA | 2 | 22 | 1 |
| 2 | No | - | ND | - | NA | 2 | 19 | 0 |
| 3 | No | - | ND | - | NA | 2 | 61 | 30 |

[1]This includes past laboratory evidence of SARS-CoV-2 infection (i.e., history of past positive SARS-CoV-2 NAAT or antigen test) or laboratory evidence of SARS-CoV-2 infection during MIS-C illness (i.e., positive SARS-CoV-2 NAAT, antigen, or anti-nucleocapsid antibody test). Spike antibody assay results are not included in this variable.
[2]Cells containing (-) indicate a negative test; ND = Not Done; Tests reported as “not done” were tests reported as not done in the medical notes or not present per our review of available medical records.

**Supplemental Table 2c.** Clinical phenotypes of three persons with illness meeting MIS-C clinical and inflammatory criteria except for a positive SARS-CoV-2 test— United States, December 2020 through August 2021

| Case No. | Clinical Phenotype[1] | Assessment by Brighton MIS-C Criteria, Level | Cardiac | | | | | Dermatologic | | Gastrointestinal | | | Hematologic | | Neurologic | | Pulmonary | | Renal |
|---|---|---|---|---|---|---|---|---|---|---|---|---|---|---|---|---|---|---|---|
| | | | Shock | ↑ Troponin | ↑ BNP/ NT-proBNP | Cardiac Dysfunction | CAA | Rash | MC Lesions | ↑ Bilirubin | ↑ AST/ALT | Diarrhea | ↑ D-dimer | ↓ Plts | Encephalopathy | Headache | Pulmonary Edema | Cough | AKI |
| 1 | D, G, R | Probable, 2b | | | | | | X | X | X | X | X | | | | | | | X |
| 2 | C, D, G, H, N | Definitive, 1 | | X | | | | | X | X | X | X | X | X | X | X | | | |
| 3 | D, G, H, N | Probable, 2a | | | | | | X | | | X | | X | | | X | | | |

[1]C=Cardiovascular (elevated troponin, elevated B-type natriuretic peptide (BNP)/ N-terminal pro hormone BNP (NT-proBNP), abnormal echocardiogram, arrhythmia); D=Dermatologic/mucocutaneous (rash, mucocutaneous lesions); G=Gastrointestinal (elevated bilirubin, elevated liver enzymes, or diarrhea); H=Hematologic (elevated D-dimer, thrombophilia, or thrombocytopenia); N=Neurologic (headache, meningismus, altered mental status); P=Pulmonary (pneumonia, acute respiratory distress syndrome); R=Renal (acute kidney injury); S=Shock (clinically noted in the medical record)

Abbreviations: CAA=coronary artery aneurysm; AKI= acute kidney injury; ↓ Plts=platelets <150,000/mcl; MC=mucocutaneous

**Supplemental Table 3.** Clinical features of 26 persons not meeting the CDC multisystem inflammatory syndrome in children (MIS-C) case definition— United States, December 2020 through August 2021

| No.[1] | Clinical Phenotype[2] | Reason for Not Meeting MIS-C Definition | Alternative Diagnosis | Laboratory Evidence of SARS-CoV-2 Infection[3] | SARS-CoV-2 Testing Results During Evaluation of Illness[4] | | |
|---|---|---|---|---|---|---|---|
| | | | | | NAAT | Anti-spike Antibody | Anti-nucleocapsid Antibody |
| 1 | D, G, R | No Positive SARS-CoV-2 Test | NA | No | - | ND | - |
| 2 | C, D, G, H, N | No Positive SARS-CoV-2 Test | NA | No | - | ND | - |
| 3 | D, G, H, N | No Positive SARS-CoV-2 Test | NA | No | - | ND | - |
| 4 | D, G, R, S | Alternative Diagnosis | Hemophagocytic Lymphohistiocytosis | No | - | + | - |
| 5 | C, D | Alternative Diagnosis | Sickle Cell Crisis | Yes | - | + | ND |
| 6 | C, D, G, H, N | Alternative Diagnosis | Murine Typhus | No | - | + | - |
| 7 | D, G, H, N | Alternative Diagnosis | Group A *Streptococcus* mediated disease | No | - | + | ND |
| 8 | G, N, P | Alternative Diagnosis | E-cigarette or Vaping-use Associated Lung Injury | No | - | ND | - |
| 9 | G, N, P | Alternative Diagnosis | Acute SARS-CoV-2 Infection | Yes | + | ND | ND |
| 10 | G, H, P | Alternative Diagnosis | Acute SARS-CoV-2 Infection | Yes | + | ND | ND |
| 11 | G, H, P | Alternative Diagnosis | Parainfluenza | No | - | ND | - |
| 12 | C, G, H | Alternative Diagnosis | Enterovirus Myopericarditis | Yes | - | + | - |
| 13 | C, N | Alternative Diagnosis | Myocarditis | No | - | ND | ND |
| 14 | C, G | Alternative Diagnosis | Myocarditis | No | - | + | - |
| 15 | C, N | Alternative Diagnosis | Myocarditis | No | - | + | ND |
| 16 | C, G | Alternative Diagnosis | Myocarditis | No | - | + | - |
| 17 | C, N | Alternative Diagnosis | Myocarditis | Yes | - | ND | + |
| 18 | C | Alternative Diagnosis | Myocarditis | No | - | ND | ND |
| 19 | C, G | Alternative Diagnosis | Myocarditis | No | - | + | ND |
| 10 | C, N | Alternative Diagnosis | Myocarditis | No | - | ND | ND |
| 21[1] | C, D, G, H, P, R, S | Alternative Diagnosis | Idiopathic cardiomyocyte necrosis | No | - | + | - |
| 22 | C, D, G, H, N, P, R | No Fever | NA | No | - | ND | ND |
| 23 | C, G, N | No Fever | NA | No | - | ND | ND |
| 24[1] | C, G, H, P, R | < 24H of Fever | NA | No | - | + | - |
| 25 | G | < 2 Organ System Involvement | NA | No | - | ND | ND |
| 26 | C, N | Inflammatory Markers Within Normal Range | NA | No | - | ND | ND |

[1]Persons numbered 21 and 24 died during illness. For person 21, medical records including autopsy report were discussed with MIS-C and immunization safety expert clinicians, physician investigators from CDC's Clinical Immunization Safety Assessment (CISA) Project including specialists in infectious diseases and cardiology, and with experts from the CDC Infectious Disease Pathology Branch who had reviewed the case independently; It was concluded that this person did not meet CDC MIS-C case definition and had an alternative diagnosis; cause of death on autopsy

was multiorgan failure due to acute heart failure due to idiopathic cardiomyocyte necrosis. Autopsy report was not available for person number 24, but this person had preexisting cardiomyopathy and did not meet MIS-C case definition because the fever criterion was not met.

[2]Clinical phenotypes are described by organ system involved and abbreviated as follows: C=Cardiovascular (elevated troponin, elevated B-type natriuretic peptide (BNP)/ N-terminal pro hormone BNP (NT-proBNP), abnormal echocardiogram, arrhythmia); D=Dermatologic/mucocutaneous (rash, mucocutaneous lesions); G=Gastrointestinal (elevated bilirubin, elevated liver enzymes, or diarrhea); H=Hematologic (elevated D-dimer, thrombophilia, or thrombocytopenia); N=Neurologic (headache, meningismus, altered mental status); P=Pulmonary (pneumonia, acute respiratory distress syndrome); R=Renal (acute kidney injury); S=Shock (clinically noted in the medical record)

[3]This includes past laboratory evidence of SARS-CoV-2 infection (i.e., past positive SARS-CoV-2 NAAT or antigen test) and evidence during MIS-C illness (i.e., positive SARS-CoV-2 NAAT, antigen, or anti-nucleocapsid antibody test); Persons numbered 5 and 12 had a positive SARS-CoV-2 NAAT test on 153 and 208 days before illness onset respectively

[4]Cells containing (-) indicate a negative test; person numbered 26 had a negative SARS-CoV-2 viral antigen test during illness. Tests reported as “not done” were tests reported as not done in the medical notes or not present per our review of available medical records.

Abbreviations: ND = Not Done; Tests reported as “not done” were tests reported as not done in the medical notes or not present per our review of available medical records; NA = Not Applicable.

**Supplemental Table 4.** Clinical phenotypes of 15 persons with multisystem inflammatory syndrome in children (MIS-C) following COVID-19 vaccination with laboratory evidence of SARS-Cov-2 infection— United States, December 2020 through August 2021

| Case No. | Clinical Phenotype[1] | Assessment by Brighton MIS-C Criteria, Level[2] | Cardiac | | | | | Dermatologic | | Gastrointestinal | | | Hematologic | | Neurologic | | Pulmonary | | Renal |
|---|---|---|---|---|---|---|---|---|---|---|---|---|---|---|---|---|---|---|---|
| | | | Shock | ↑ Troponin | ↑ BNP/NT-proBNP | Cardiac Dysfunction | CAA | Rash | MC Lesions | ↑ Bilirubin | ↑ AST/ALT | Diarrhea | ↑ D-dimer | ↓ Plts | Encephalopathy | Headache | Pulmonary Edema | Cough | AKI |
| 1 | C, D, G, H, N, P, R, S | Definitive, 1 | X | X | X | | | | X | X | | X | X | | X | | X | X | X |
| 2 | C, D, G, N | Definitive, 1 | | X | X | | | X | | | | X | | | | X | | | |
| 3 | C, D, G, H, P, R, S | Probable, 2b | X | X | X | X | | X | X | X | X | X | X | | | | X | | X |
| 4 | C, D, G, N | Probable, 2a | | | X | | | | X | | | X | X | | | X | | | |
| 5 | C, G, H | Definitive, 1 | | X | X | X | X | | | | X | | X | | | | | | |
| 6 | C, G, H, R, S | Definitive, 1 | X | X | X | X | | | | X | | | | X | | | | | X |
| 7 | G, H, N | Not a case, 5 | | | | | | | | | X | | X | | | X | | | |
| 8 | C, G, H, N, P | Definitive, 1 | | X | | X | | | | X | X | X | | X | | X | | X | |
| 9 | C, D, G, N, P | Definitive, 1 | | | | | X | X | X | | X | X | | | | X | | X | |
| 10 | C, H, P, S | Not a case, 5 | X | | X | | | | | | | | X | | | | | X | |
| 11 | C, D, G, H, N, P | Definitive, 1 | | X | | | | X | | | X | X | X | X | | X | | X | |
| 12 | C, D, G, H, N, P, S | Definitive, 1 | X | X | X | X | | X | X | | | X | | X | | X | X | | |
| 13 | C, D, G, H, P, S | Definitive, 1 | X | X | | X | | X | | X | | | X | | | | | X | |
| 14 | G, H, N | Not a case, 5 | | | | | | | | X | X | | | X | | X | | | |
| 15 | C, G, H, N, P, R, S | Definitive, 1 | X | X | X | X | | | | X | X | | X | | | X | X | | X |

[1]C=Cardiovascular (elevated troponin, elevated B-type natriuretic peptide (BNP)/ N-terminal pro hormone BNP (NT-proBNP), abnormal echocardiogram, arrhythmia); D=Dermatologic/mucocutaneous (rash, mucocutaneous lesions); G=Gastrointestinal (elevated bilirubin, elevated liver enzymes, or diarrhea); H=Hematologic (elevated D-dimer, thrombophilia, or thrombocytopenia); N=Neurologic (headache, meningismus, altered mental status); P=Pulmonary (pneumonia, acute respiratory distress syndrome); R=Renal (acute kidney injury); S=Shock (clinically noted in the medical record)

[2]Three persons were considered not a case per Brighton case definition: two had <2 clinical features and one lacked evidence of disease activity

Abbreviations: CAA=coronary artery aneurysm; AKI= acute kidney injury; ↓ Plts=platelets <150,000/mcl; MC=mucocutaneous

**Supplemental Table 5.** Clinical phenotypes of six persons with multisystem inflammatory syndrome in children (MIS-C) following COVID-19 vaccination without laboratory evidence of SARS-Cov-2 infection— United States, December 2020 through August 2021

| Case No. | Clinical Phenotype[1] | Assessment by Brighton MIS-C Criteria, Level | Cardiac | | | | | Dermatologic | | Gastrointestinal | | | Hematologic | | Neurologic | | Pulmonary | | Renal |
|---|---|---|---|---|---|---|---|---|---|---|---|---|---|---|---|---|---|---|---|
| | | | Shock | ↑ Troponin | ↑ BNP/ NT-proBNP | Cardiac Dysfunction | CAA | Rash | MC Lesions | ↑ Bilirubin | ↑ AST/ALT | Diarrhea | ↑ D-dimer | ↓ Plts | Encephalopathy | Headache | Pulmonary Edema | Cough | AKI |
| 16 | C, H, N, P | Definitive, 1 | | | X | | | | | | | | X | | | X | | X | |
| 17 | C, D, G, H, P, S | Definitive, 1 | X | X | | | | X | | X | X | | X | X | | | | X | |
| 18 | C, D, H | Definitive, 1 | | X | X | | | X | X | | | | X | | | | | | |
| 19 | C, D, G, H, N | Definitive, 1 | | X | | | | | X | | | X | X | | | X | | | |
| 20 | C, D, G, H, P, R | Definitive, 1 | X | X | X | X | | X | | | | X | X | | | | X | X | X |
| 21 | C, G, N | Probable, 2b | | X | | | | | | | | X | | | X | X | | | |

[1]C=Cardiovascular (elevated troponin, elevated B-type natriuretic peptide (BNP)/ N-terminal pro hormone BNP (NT-proBNP), abnormal echocardiogram, arrhythmia); D=Dermatologic/mucocutaneous (rash, mucocutaneous lesions); G=Gastrointestinal (elevated bilirubin, elevated liver enzymes, or diarrhea); H=Hematologic (elevated D-dimer, thrombophilia, or thrombocytopenia); N=Neurologic (headache, meningismus, altered mental status); P=Pulmonary (pneumonia, acute respiratory distress syndrome); R=Renal (acute kidney injury); S=Shock (clinically noted in the medical record)

Abbreviations: CAA=coronary artery aneurysm; AKI= acute kidney injury; ↓ Plts=platelets <150,000/mcl; MC=mucocutaneous

**Supplemental Figure 1**. Number of MIS-C cases with laboratory evidence of SARS-CoV-2 infection, by days from COVID-19 vaccine— United States, December 2020 through August 2021, n=15. A. Number of cases by number of days from last COVID-19 vaccine dose to onset of MIS-C B. Number of cases by number of days from last COVID-19 vaccine dose to hospital admission. C. Number of cases by number of days from Dose 1 of COVID-19 vaccine to onset of MIS-C.

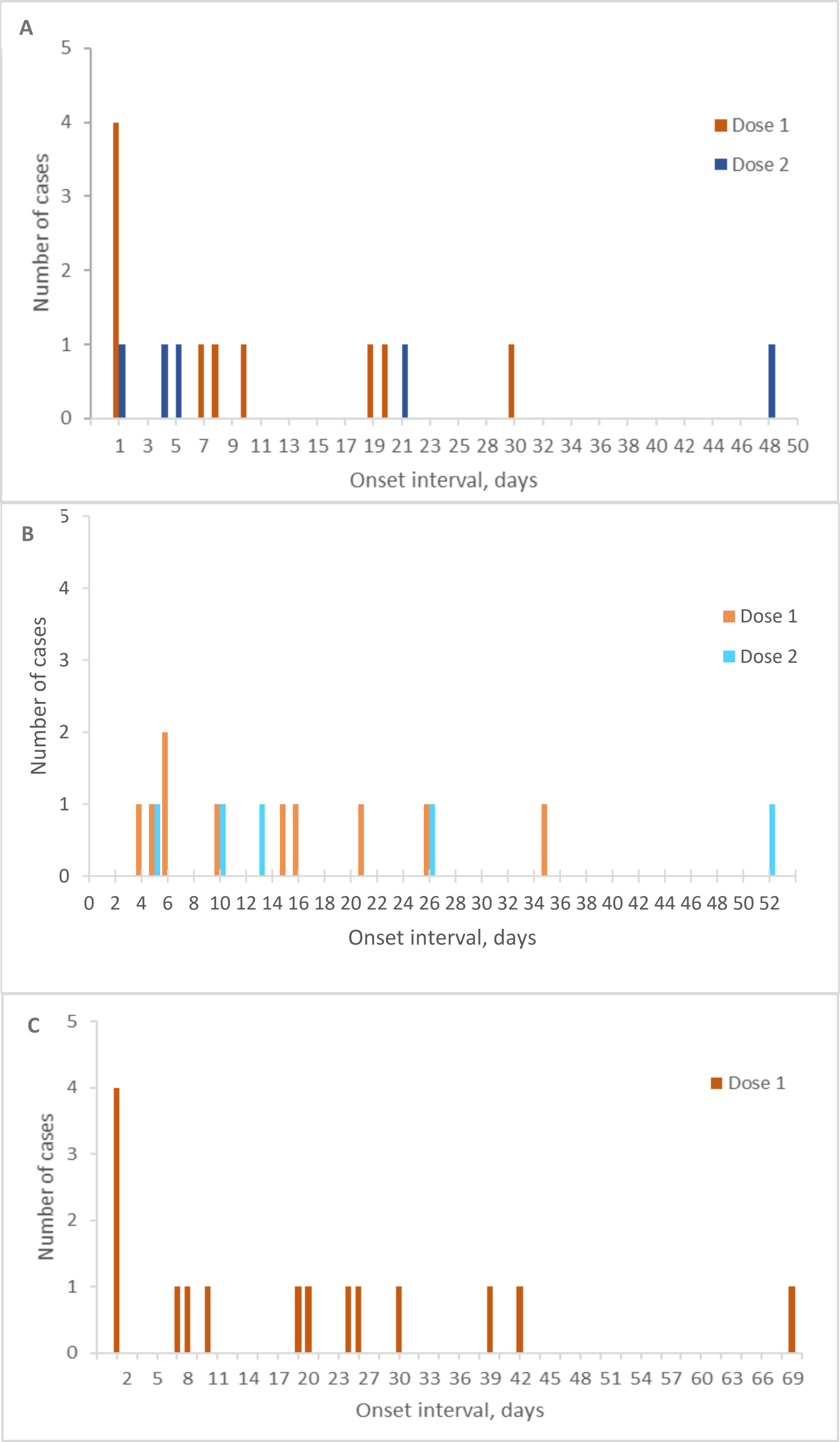

**Supplemental Figure 2.** Number of MIS-C cases without laboratory evidence of SARS-CoV-2 infection (solid bars, n=6) and those with illness meeting MIS-C clinical and inflammatory criteria except for a positive SARS-CoV-2 test (hashed bars, n=3), by days from COVID-19 vaccine— United States, December 2020 through August 2021 A. Number of patients by number of days from last COVID-19 vaccine dose to onset of MIS-C. B. Number of patients by number of days from last COVID-19 vaccine dose to hospital admission. C. Number of patients by number of days from Dose 1 of COVID-19 vaccine to onset of MIS-C. Note break in x-axes.

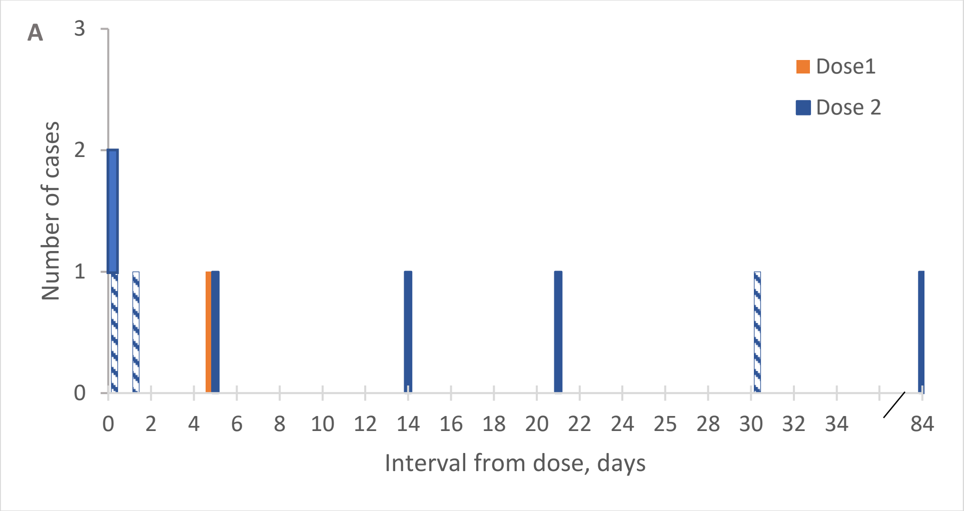


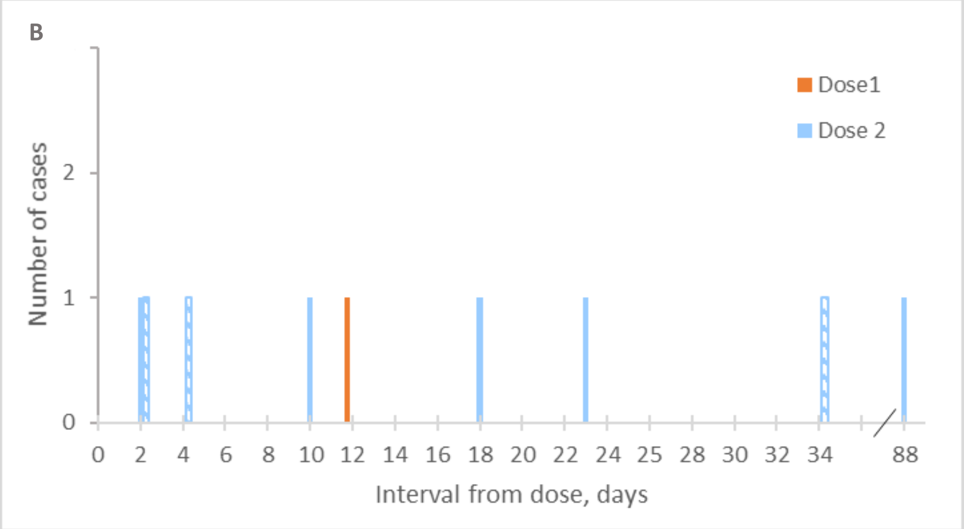


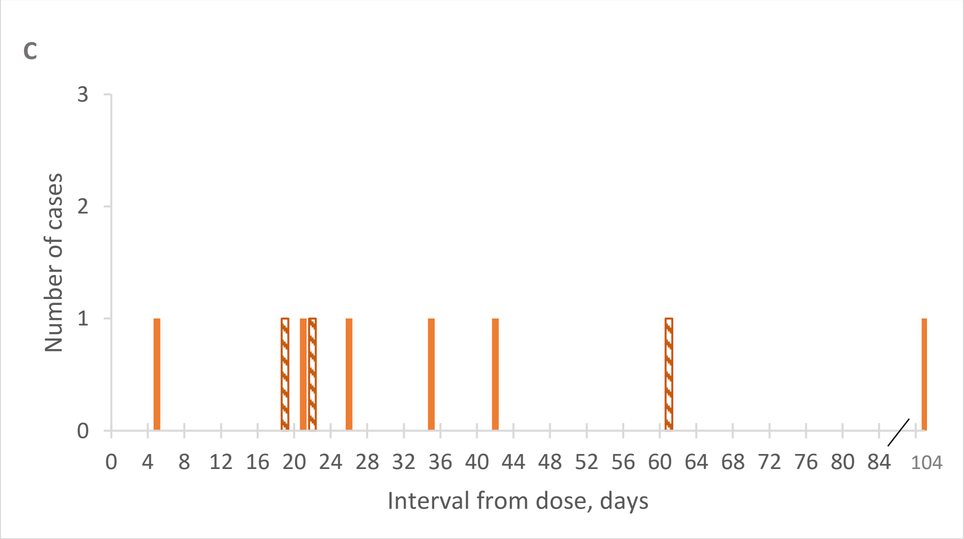
